# Generative embedding of sparse data with a tabular foundation model for dengue anticipatory action: a machine learning approach

**DOI:** 10.64898/2026.07.03.26357228

**Authors:** Keanu John Pelitro, Julia Fye Manzano, Troy Owen Matavia, Kylone Soriano, Klara Bilbao, Gereka Marie Garcia, Aira Joy Delos Angeles, Alfredo Mahar Lagmay, DJ Darwin Bandoy

**Affiliations:** University of the Philippines Resilience Institute, Quezon City, Philippines; College of Veterinary Medicine, University of the Philippines, Los Banos, Laguna, Philippines; National Institute of Geological Sciences, University of the Philippines Diliman, Quezon City, Philippines

## Abstract

**Background:** Early outbreak detection has largely relied on complex, data-intensive models with limited applicability to low-resource surveillance. Even state-of-the-art tabular foundation models require dense datasets for fine-tuning to capture disease transmission dynamics. We address this by building a domain-mechanistic generative embedding from cases and rainfall to detect early epidemic onset.

**Methods:** We build a generative, domain-mechanistic embedding from sparse case and rainfall data into 132 features, converting limited inputs into a structured representation of transmission for outbreak-onset detection. A tabular foundation model was evaluated by leave-one-year-out validation with cluster-bootstrap intervals across 17 Philippine regions and eight dengue-endemic countries, benchmarked against raw data columns and catch22.

**Findings:** Raw columns used as input to the tabular foundation model were weakly predictive of dengue outbreak onset (AUROC 0·56–0·70). The generative embedding improved detection to 0·77 across countries and 0·89 across regions (+0·205 and +0·183; paired cluster-bootstrap p≤0·006). Calibration error was lower at the regional scale than at the country scale (expected calibration error 0·067 and 0·149). Strongly seasonal regions and countries were the most predictable (Philippine Type I region mean 0·87; Mexico 0·94, Brazil 0·93, the Philippines 0·91), whereas countries with year-round or coastally opposing rainfall were weaker or below chance (Singapore 0·69, Sri Lanka 0·42), and countries left with only one or two seasons after applying the onset rule gave unreliable estimates.

**Interpretation:** Under sparse surveillance conditions, predictive capacity depended strongly on the representation supplied to the tabular foundation model. The generative embedding translates climate and epidemiological variables into actionable early-warning signals by capturing underlying transmission mechanisms, whose accuracy scales with local seasonal dynamics. This approach provides a viable pathway for extending prospective outbreak surveillance in data-limited settings, and indicates that mechanism-grounded embeddings could calibrate transmission-acceleration models at aggregated scales to improve their predictions.

**Funding:** National Institute of Environmental Health Sciences, National Institutes of Health (award P20ES036118).

**Research in context:** *Evidence before this study:* We searched PubMed, Web of Science, and Google Scholar up to April 2026, combining “dengue” with “early warning”, “outbreak prediction”, or “forecasting”, and “machine learning”, “foundation model”, or “feature engineering”. Foundation models have recently been applied to epidemic modeling across pathogens, including dengue, forecasting incidence from raw surveillance series with minimal retraining. Their main limitation is the absence of a disease transmission mechanism in an otherwise black-box model. Closing that gap has been framed as a problem of scale, requiring large-scale epidemic-specific datasets for retraining.

*Added value of this study:* We show that for sparse dengue surveillance, predictive capacity is set by how the input represents transmission dynamics, and that the transmission mechanism absent from a foundation model can be supplied through a generative embedding. Computing 132 features from these quantities lifted the tabular foundation model from near-chance and weak detection on the raw columns at the aggregated country and regional scales (AUROC 0·56 and 0·70) to stronger detection of the epidemic onset phase, with the clearest operational performance at the regional scale (AUROC 0·89). Regional scores detected epidemic onset, but calibration slopes below one showed that absolute probabilities require local recalibration before use. As climate change outpaces the build-out of surveillance, the capacity to anticipate outbreaks from counts and weather alone may matter most where data are limited.

*Implications of all the available evidence:* Recent benchmarks proposed adding disease mechanisms to foundation models through fine-tuning. A generative embedding of the transmission mechanism achieved strong detection performance on sparse data with no model retraining. The regional scale is the appropriate calibration and deployment scale for dengue anticipatory action. Moreover, the country scale is informative as a transportability test because heterogeneous reporting systems, asynchronous climate zones, and sparse retained seasons can weaken or invalidate national estimates. This principle, reconstructing a mechanism-grounded representation so a general-purpose model can process transmission, could extend to other climate-sensitive diseases that record little more than counts and weather.

## Introduction

Dengue early-warning systems aim to trigger appropriate and timely response before transmission escalates.^1^ WHO’s Early Warning and Response System, for instance, ties meteorological alarm indicators to case thresholds, and Bayesian spatiotemporal models forecast incidence using climatic factors, but these approaches require sufficient data to be effective.^2^ As a response to sparse epidemiological data, foundation models are reshaping how dengue case time series are analyzed. TabPFN, a tabular foundation model, makes accurate predictions on new, small datasets through approximate Bayesian inference and often matches or surpasses models tuned from scratch.^3,4^ Applied to infectious disease, foundation models have been used to forecast incidence and the timing of seasonal peaks directly from case counts.^5–7^

While generating robust predictions, a key limitation is that foundation models carry no representation of how diseases are transmitted. Classical epidemic models encode susceptible depletion, serial intervals, and reproductive number as transmission parameters, whereas a general-purpose foundation model reads a time series dataset with no such structure.^1,5,7^ The state-of-the-art approach for this missing mechanism is to fine-tune or retrain foundation models on large volumes of epidemic data, so that the transmission structure is learned as a prior, something that is not available in resource-constrained settings.^5,7^ High-quality records are scarce and heavily under-reported in endemic regions, which limits the interpretability and utility of tabular foundation models. ^8–10^

Fine-tuning disease mechanisms into a tabular foundation model requires high-dimensional data. This requirement limits use in data-constrained settings. A mechanistic model addresses this by defining a feature space in which transmission is explicit, so a classifier reads structure that the raw series leaves hidden.^11^ Generative embedding works on this principle, taking the inferred parameters of a fitted model as features and improving accuracy and interpretability under small samples.^12,13^ The principle depends on fitting a model, and fitting depends on dense data. We adapt the approach by encoding each transmission relationship as a deterministic transform of cases and rainfall, applied directly to the series. The features are computed, not learned, so the representation requires none of the data that fitting would demand. Rainfall rises before transmission where dengue is climate-paced, so the transformations expose the acceleration of cases and the phase of the seasonal cycle,^14^ which rise before incidence as transmission approaches a tipping point.^15–18^ Each transform holds at any record length, so a short record carries the representation and the foundation model detects epidemic onset.^16,17^

We hypothesize that generative embedding provides mechanistic transmission representations to a tabular foundation model, enabling accurate and calibrated detection of dengue epidemic onset from sparse weekly case counts and rainfall. We evaluate outbreak onset detection by leave-one-year-out validation across 17 Philippine regions and eight dengue-endemic countries. We first determine whether the embedding makes onset detectable from the two raw time-series columns. We then trace improvement to the generated transmission features, using catch22 as a generic-shape control and permutation importance to rank what detects onset earliest. We mapped performance across climate types. Finally, we assess calibration, testing whether the probabilities can be acted on without rescaling. These analyses determine whether mechanistic representation can render sparse counts and rainfall actionable for dengue early warning.

## Methods

### Data sources and analytical scales

Our analysis spanned two nested geographic scales (table 1). The regional scale was weekly counts for the 17 Philippine administrative regions over 2016–2025, labelled by climate type.^19^ The national scale used OpenDengue counts^20^ for eight dengue-endemic countries (Brazil, Colombia, Mexico, Peru, the Philippines, Singapore, Sri Lanka, and Taiwan), each paired with a weekly rainfall series from NASA rainfall data. Every observation was dated to the Monday of its ISO epidemiological week. A year was evaluable when its series ran long enough to define the target and fell outside the excluded seasons: the COVID-19-disrupted years 2020 and 2021.

**Table 1:** Evaluation scales, data sources, and the two raw input columns.

| Scale | Units | Years | Raw inputs | Source |
| --- | --- | --- | --- | --- |
| Regional | 17 Philippine regions | 2016–2025 | weekly cases, weekly rainfall | Philippine DOH / Humanitarian Data Exchange |
| Country | 8 endemic countries | 2016–2025* | weekly cases, weekly rainfall | OpenDengue |

*Target: the epidemic onset phase of each season. Onset = the first week the constant transmission-acceleration (STA/LTA) detector fires inside the anticipatory metrics framework, using a 4-week short-term over 26-week long-term case mean, 2-week guard, activation 1.33, deactivation 0.73, calendar-year resets, run on the full continuous per-unit series (the excluded 2020–21 seasons feed the long-term mean, then are dropped at evaluation).<sup>17</sup> The epidemic onset phase is every week from onset up to, but not including, the peak; all other weeks are negatives. Features use information up to week $t$ only. \*Country analysis used available OpenDengue weeks within 2016–2025 after excluding 2020–2021 from evaluation and dropping seasons without a qualifying onset.*

### Target: epidemic onset

Yearly, we set the annual peak as the week of maximum cases and the onset as the first transmission-acceleration trigger before that peak. A trigger qualified only when it fell inside the season’s anticipatory window: the actionable weeks four to eight weeks before the peak, or the smallest block of weeks around the peak that held at least 70% of the season’s cases.^17,21^ A season with no qualifying trigger before its peak had no defined onset and was dropped. Weeks from onset to the week before the peak, the epidemic onset phase, were labelled positive, and all other weeks of a retained season were labelled negative. Each retained year was scored in full. This target is defined retrospectively from each retained season; prospective deployment would require calibration.

### Generative embedding of features

Generative embedding maps raw data through a generative model into a feature space that the foundation model reads. We adapt and extend this approach by reconstructing the transmission process from case and rainfall series as deterministic features read by a tabular foundation model.^12^ We expanded the two raw columns into 132 features: 108 domain features (63 rainfall-derived, 38 case-derived, and seven calendar) and 24 canonical time-series features as a control (table S4). Multi-scale case and rainfall views capture recent transmission and the delayed effect of rainfall on vector habitat through lagged values, rolling means, standard deviations, and extrema.^14,22^ Rainfall-accumulation windows encode habitat persistence and the breeding-threshold response, and momentum terms encode the rate of change. The transmission-acceleration feature is a short-term to long-term average ratio adapted from seismic event detection, with a four-week short-term and a 26-week long-term window separated by a guard band.^18,19^ Seasonality enters through a prior-year’s week-of-year climatology and Fourier terms; rainfall accumulation through logistic phase-transition views and a saturating Hill term that expresses the threshold and saturation behaviour of mosquito habitat; and low-frequency structure through 26-week and 52-week anomalies and a wet-season run length that stands in for El Niño–Southern Oscillation forcing without an external index.^14^ Critical-slowing-down indicators, including rolling variance and lag-one autocorrelation, follow the early-warning theory of critical transitions.^16,23^

A susceptibility reconstruction class encodes the multi-year susceptibility rhythm causally within each unit: the inter-epidemic interval since incidence last reached the expanding 90th-percentile epidemic level, a 52-week cumulative burden, and a susceptibility proxy combining a long interval with low recent burden.^24^ A unit test confirms that perturbing future case values leaves every earlier-week feature unchanged, which verifies the causal construction directly in code. The canonical control is the catch22 set, computed causally over a trailing 52-week window of both series.^25^

### Model, comparator, and evaluation

Detection was evaluated with the tabular foundation model TabPFN, run on a CPU with no model-weight updating or gradient retraining on the surveillance data. Validation was expanding leave-one-year-out: for each evaluable year, the model received all prior evaluable years as its in-context training set and was scored on the held-out year. Preprocessing was fitted on the training rows only (median imputation then a rank-Gaussian transform), with isotonic recalibration fit on training-fold predictions.^26^ The in-context training set was capped while retaining both classes. The full feature set was retained a priori so that the evaluation tested the complete representation rather than a fold-specific selected subset.^26^

We report AUROC, AUPRC, the Brier score, and the expected calibration error (with 95% confidence intervals), with feature attribution by leave-one-year-out permutation importance. Uncertainty and between-representation comparisons were obtained from a paired cluster bootstrap with 2000 resamples that resampled whole unit-years, the natural exchangeable block under leave-one-year-out, so that 95% confidence intervals and p-values respected the temporal clustering of weekly observations; the effective sample size is the number of unit-years (89 across regions and 32 across countries). Because every representation was scored on the identical held-out weeks, representations were compared as paired contrasts. One comparison, the reconstructed representation against the two raw columns in AUROC, was pre-specified as confirmatory; all other contrasts were treated as exploratory with Benjamini–Hochberg control of the false-discovery rate, and the DeLong test was reported as a secondary check on paired AUROC differences. Calibration was assessed by the Cox calibration slope and intercept and the Spiegelhalter z test alongside the Brier score and expected calibration error, each with cluster-bootstrap intervals. Per-region and per-country detection was judged against chance by whether the lower bound of the bootstrap interval exceeded 0·5, and stability to the model’s internal randomness was checked by repeating the fit across random seeds. A rainfall-trigger analysis derived, within fold, the rainfall representation and lead time that best anticipate onset. The rainfall-dynamics and accumulation representations were then contrasted by the same paired cluster bootstrap on the weeks both could score.

### Reproducibility, ethics, and reporting

The study is reported following TRIPOD+AI^27^, the reporting standard for prediction-model studies that use machine learning, and the completed checklist accompanies the submission. The analytical code and the deidentified weekly datasets are available as described in the data sharing statement. The study was approved by the University of the Philippines Research Ethics Committee (Protocol 2024-0004-F-FMDS). Informed consent was waived, as is standard for retrospective analysis of anonymised public surveillance data, because every record was an aggregated weekly total carrying no individual-level identifier.

## Results

### Generative embedding enhances tabular foundation model performance

Reading case counts and rainfall directly, the tabular foundation model (TabPFN) detected no onset signal across countries and only a weak one across regions (AUROC 0·562, 0·704; figure 1A, table 2). The generative embedding raised detection at both scales, to AUROC 0·767 across countries and 0·887 across regions. TabPFN gained AUROC +0·205 and +0·183 with 95% confidence intervals excluding zero (paired cluster-bootstrap p=0·006 and p<0·001; table 3). Once the raw series were reconstructed into the embedding, the model became substantially better at detecting the epidemic onset phase (base rate 0·261 country, 0·195 regional; AUPRC rose from 0·32 to 0·52 across countries and 0·32 to 0·67 across regions; table 2). The domain-agnostic catalogue catch22 also raised detection above the raw inputs, to 0·741 across countries and 0·837 across regions (table 2), confirming that the onset signal is readable once the raw series is re-expressed in a richer feature space. The generative embedding matched or exceeded catch22 at every scale, consistent with the transmission mechanism encoded by the domain features.

**Figure 1:**
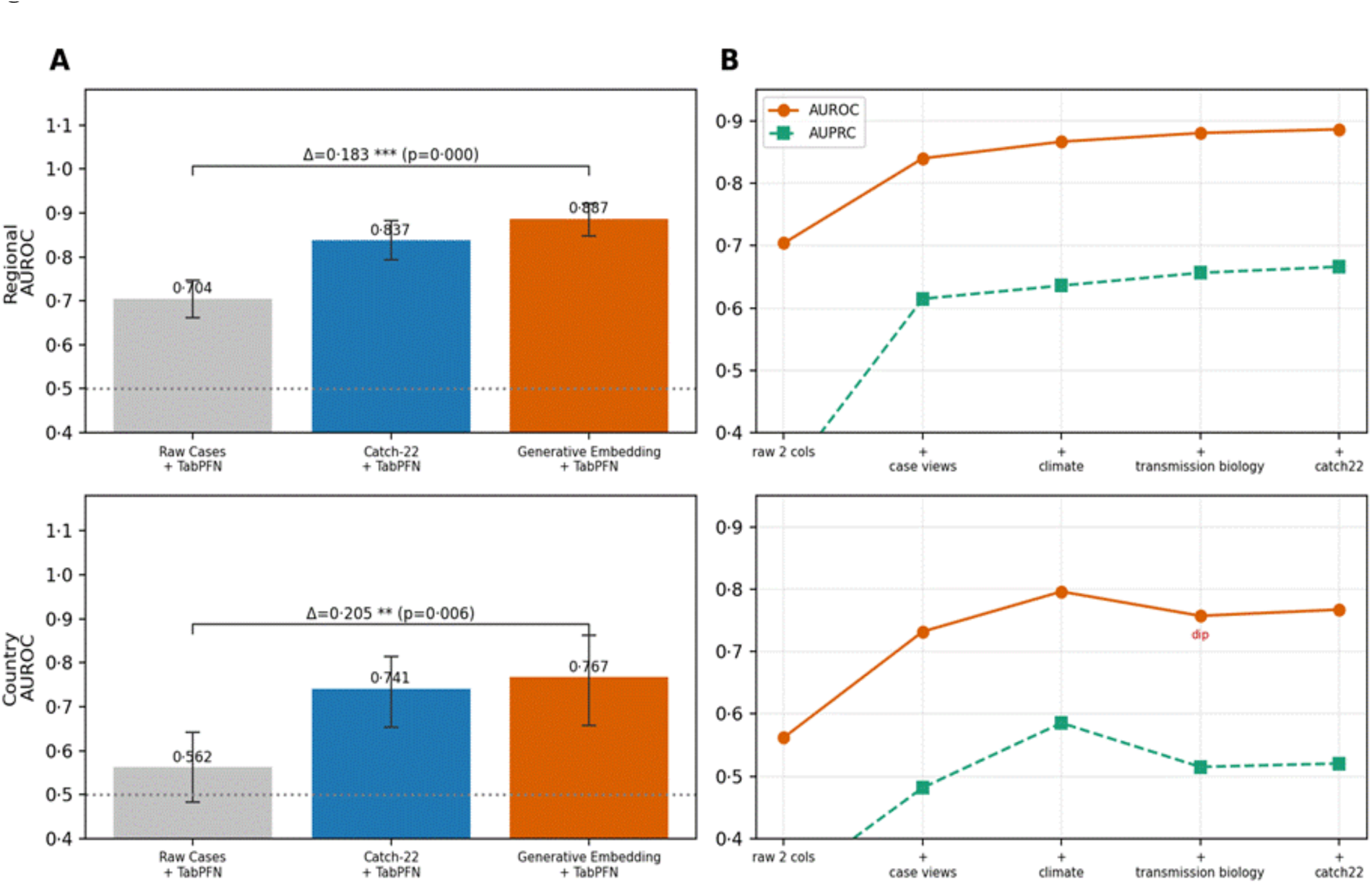
Representation governs early-warning detection. Rows are the two scales (regional, country). (A) Leave-one-year-out AUROC for three representations, each read by the same tabular foundation model: the raw two-column table (Raw Cases + TabPFN), the generic canonical catch22 representation (Catch-22 + TabPFN), and the reconstructed embedding (Generative Embedding + TabPFN). Error bars are 95% cluster-bootstrap confidence intervals (resampling whole unit-years); the bracket gives the Generative-Embedding-minus-Raw-Cases AUROC difference with its bootstrap p value. The embedding beats raw cases at both scales. (B) Cumulative enrichment ladder; the generic catch22 rung adds only slightly at the regional and country scales.

**Table 2:** **Early-warning detection by representation and scale** (expanding leave-one-year-out tabular foundation model; epidemic onset phase target defined in Table 1). AUROC and AUPRC are point estimates with 95% cluster-bootstrap confidence intervals.

| Scale | Representation | n | AUROC (95% CI) | AUPRC (95% CI) | Brier |
| --- | --- | --- | --- | --- | --- |
| Regional | Raw Cases + TabPFN | 2 | 0.704 (0.662–0.748) | 0.322 (0.292–0.357) | 0.232 |
|  | Catch-22 + TabPFN | 26 | 0.837 (0.792–0.882) | 0.530 (0.474–0.597) | 0.133 |
|  | Generative Embedding + TabPFN | 132 | 0.887 (0.847–0.922) | 0.666 (0.605–0.729) | 0.104 |
| Country | Raw Cases + TabPFN | 2 | 0.562 (0.483–0.643) | 0.319 (0.252–0.396) | 0.252 |
|  | Catch-22 + TabPFN | 26 | 0.741 (0.653–0.814) | 0.459 (0.355–0.559) | 0.190 |
|  | Generative Embedding + TabPFN | 132 | 0.767 (0.658–0.863) | 0.520 (0.363–0.691) | 0.188 |

*Test weeks: regional 4625, country 1656; epidemic onset phase base rate 0.195, 0.261; year-clusters 89, 32. The Generative Embedding representation is 108 domain features plus 24 generic canonical (catch22) features; Catch-22 pairs the two raw columns with the 24 catch22 features. All estimates use $B = 2000$ cluster-bootstrap resamples.*

**Table 3:** Statistical comparison of representations (paired cluster bootstrap).

| Scale | ΔAUROC (95% CI) | ΔAUPRC (95% CI) | ΔBrier (95% CI) | Bootstrap p | DeLong z (p) |
| --- | --- | --- | --- | --- | --- |
| Regional | +0.183 (0.138–0.226) | +0.345 (0.274–0.415) | −0.128 (−0.149 to −0.106) | <0.001 | −19.94 (<0.0001) |
| Country | +0.205 (0.067–0.328) | +0.202 (0.045–0.368) | −0.064 (−0.122 to −0.004) | 0.006 | −9.79 (<0.0001) |

*Positive ΔAUROC/ $\Delta$ AUPRC and negative $\Delta$ Brier favour the Generative Embedding. The confirmatory enrichment over raw inputs is significant at the regional and country scales by both the cluster bootstrap and DeLong. No external forecasting comparator is included.*

### Generative embedding makes the foundation model’s predictions interpretable

To make TabPFN’s predictions interpretable, we scored cumulative versions of the generative embedding, so each AUROC gain could be read from the generated feature block added at that step. Starting from raw cases and rainfall, case-derived features raised detection to AUROC 0·840 across regions and 0·732 across countries (figure 1B; table S5). Climate and rainfall-accumulation features raised AUROC further to 0·867 and 0·796, placing additional signal in the rainfall-climate part of the embedding (figure 1B; table S5). Transmission-biology features produced domain-only AUROC of 0·881 across regions and 0·757 across countries (table S5). catch22 was added as the generic time-series control and changed AUROC by +0·006 across regions and +0·010 across countries (table S2). The improvement in predictability remained readable within the generated representation.

### Embedding ranks onset detection features

Out-of-fold permutation importance ranked all 132 generated and control features as candidate onset detectors. At the regional scale, the strongest features were acceleration-derived: the transmission-acceleration ratio (ΔAUROC 0·112), its interaction with recent rainfall (0·047), and its week-on-week change (0·032; figure 2A; table S6). Other features followed, including rainfall-season interactions, climate anomaly terms, seasonal phase, and susceptibility reconstruction features. Across countries, lag-one case autocorrelation ranked first (0·049), followed by the transmission-acceleration ratio (0·045), its week-on-week change (0·030), and its rainfall interaction (0·023; figure 2B; table S6). The feature search produced a compact set of onset predictors from the embedding, with acceleration detectors ranking among the strongest at every scale and catch22 features contributing mainly generic temporal dependence.

**Figure 2:**
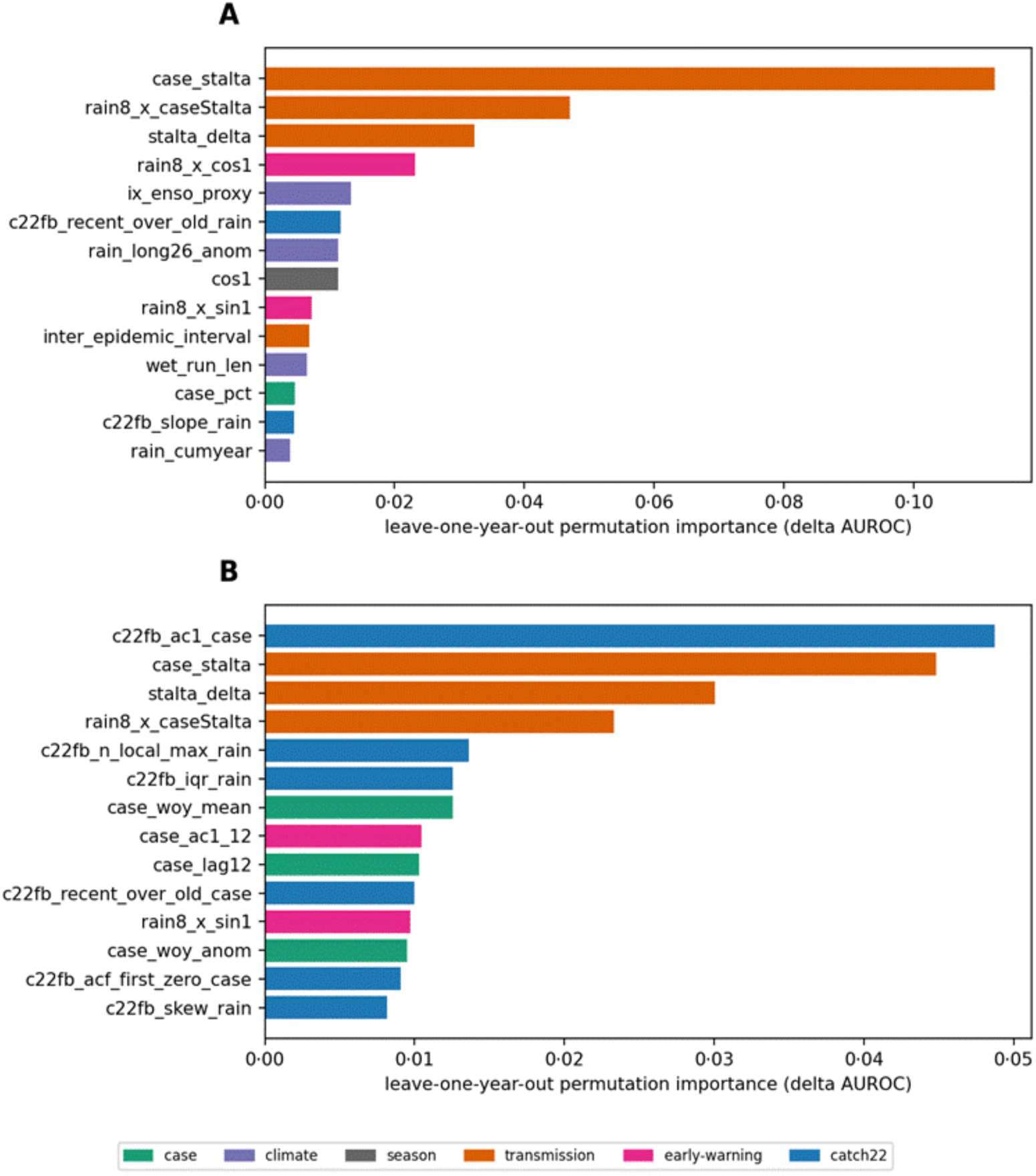
Feature attribution by scale. Horizontal bars show the leave-one-year-out permutation importance (loss in AUROC on permuting the feature) for the 14 most predictive features at each scale, coloured by group. Recent case dynamics, temporal dependence, and transmission-acceleration coordinates dominate; rainfall accumulation and seasonality contribute. Generic canonical (catch22) features also capture temporal structure, especially at the country scale.

### Predictability scales with the climate type

Performance depends on how strongly a setting’s transmission is paced by rainfall. Testing detection by region and grouping by PAGASA climate type, detection varied with the strength of rainfall seasonality (figure 3; table S7). Type I regions reached the highest mean AUROC at 0·87, led by Ilocos Region (Region I) at 0·97, Central Luzon (Region III) at 0·96, and Western Visayas (Region VI) at 0·96. The other types were lower and more variable, with mean AUROC of 0·65 for Type II, 0·85 for Type III, and 0·77 for Type IV. The across-type difference was not significant (Kruskal–Wallis H=5·99, p=0·112; 17 regions across four types; table S7). By the 95% interval, AUROC did not exceed chance in three regions: CALABARZON (Region IV-A), Eastern Visayas (Region VIII), and SOCCSKSARGEN (Region XII) (table S1).

**Figure 3:**
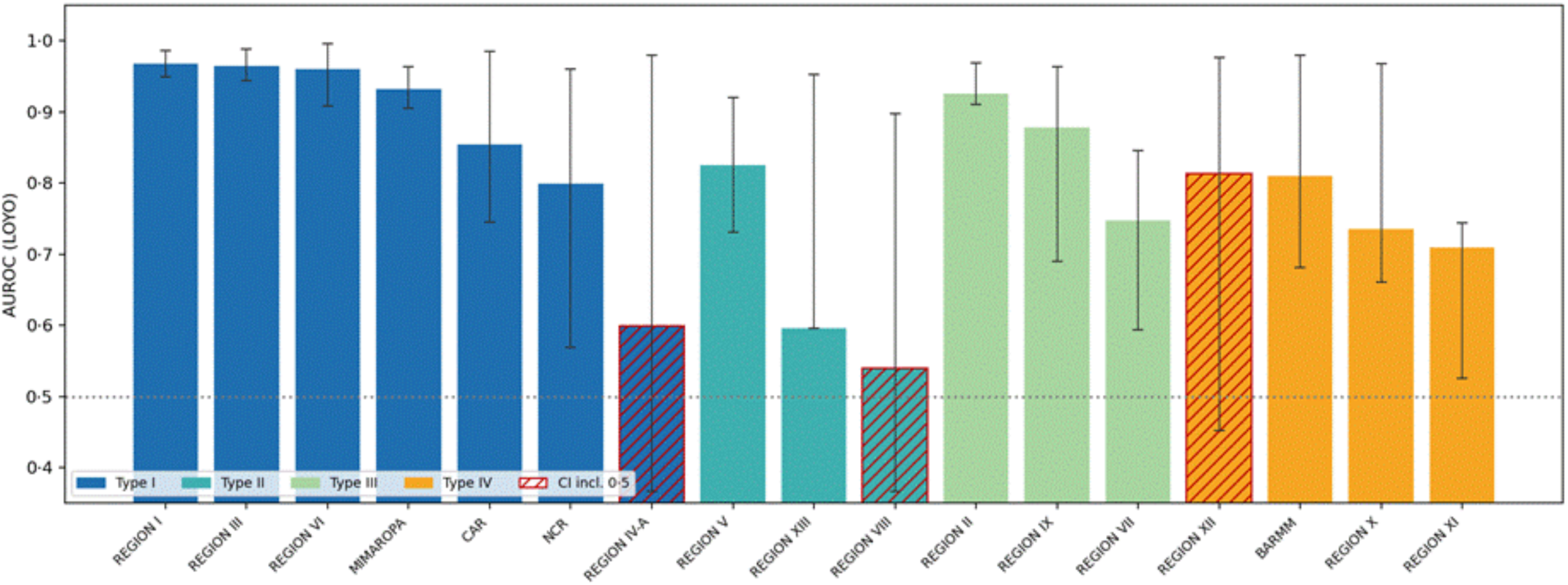
Regional predictability tracks the climate regime. Leave-one-year-out AUROC for each of the 17 Philippine regions, coloured by PAGASA climate type, with 95% cluster-bootstrap intervals over years; regions whose interval includes 0·5 are hatched. Mean detection is highest in the strongly seasonal Type I regions; the across-type difference is not significant (Kruskal-Wallis H = 5·99, p = 0·112). The dotted line marks chance; 14 of 17 regions clear chance. Some regions have wide or degenerate intervals because the no-fallback rule leaves them few evaluable seasons.

The strongly seasonal countries were also the most predictable, with Mexico at 0·94, Brazil at 0·93, and the Philippines at 0·91 (table 4). Systems with year-round or coastally opposing seasonality were weaker, including Peru at 0·75, Singapore at 0·69, and Sri Lanka at 0·42. By the 95% interval, Singapore and Sri Lanka’s AUROC did not exceed chance (table 4). Colombia posted the highest AUROC, 0·977, but with a zero-width confidence interval (0·977–0·977) that marks only a single evaluable season, and this near-perfect value reflects lack of data, not robust detection. Its Pacific and Caribbean coasts have opposing rainfall seasons that can cancel the national seasonal signal, so the acceleration detector identified a qualifying onset in only one season.^28^

**Table 4:** Per-country detection at the national scale (Generative Embedding + TabPFN, leave-one-year-out). Country-level analysis tests transportability across national surveillance systems; n is the number of evaluated weeks after dropping seasons with no detectable onset.

| Country | AUROC (95% CI) | n weeks |
| --- | --- | --- |
| Colombia | 0·977 (0·977–0·977) | 52* |
| Mexico | 0·938 (0·923–0·964) | 260 |
| Peru | 0·749 (0·710–0·759) | 104 |
| Brazil | 0·934 (0·838–0·990) | 156 |
| Philippines | 0·907 (0·896–0·957) | 149 |
| Singapore | 0·693 (0·253–0·912) | 208 |
| Taiwan | 0·760 (0·667–0·872) | 104* |
| Sri Lanka | 0·417 (0·112–0·742) | 259 |

*By interval status, six of eight country estimates exceeded chance performance ( $AUROC > 0.5$ ), but interpretability depended on the number and coherence of retained seasons. Countries marked with an asterisk (\*) are boundary cases in which only one or two seasons remained after applying the onset rule, yielding unreliable estimates that should not be interpreted as robust national validation.*

Reconstructed climate features have an anchoring structure where rainfall has a clear season. Where rainfall lacks seasonality, the rainfall-paced component of dengue transmission is weaker or obscured, and climate-based reconstruction cannot recover a seasonal signal that is absent or aggregated away in the data.

### Risk scores stratify onset at the operational scale

At the regional scale, the embedding produced a clear risk gradient. Non-onset weeks clustered at low scores, onset weeks shifted toward high scores, and the reliability curve stayed close to the diagonal, with an expected calibration error of 0·067 (figure 4; table S9). This pattern extends the detection results, where regional detection reached AUROC 0·887 and AUPRC 0·666. At the country scale, the pooled probability scale was less stable. When national surveillance systems with different reporting practices, base rates, and seasonal structures were combined, the score distributions overlapped more and expected calibration error rose to 0·149. Calibration slopes below one at both scales showed that the raw scores were too extreme as absolute probabilities. However, the regional ranking remained the actionable signal because the ordering of weeks by predicted risk was preserved. Local recalibration can map these onset-risk strata onto response thresholds and pre-position vector-control capacity ahead of the expected dengue peak.

**Figure 4:**
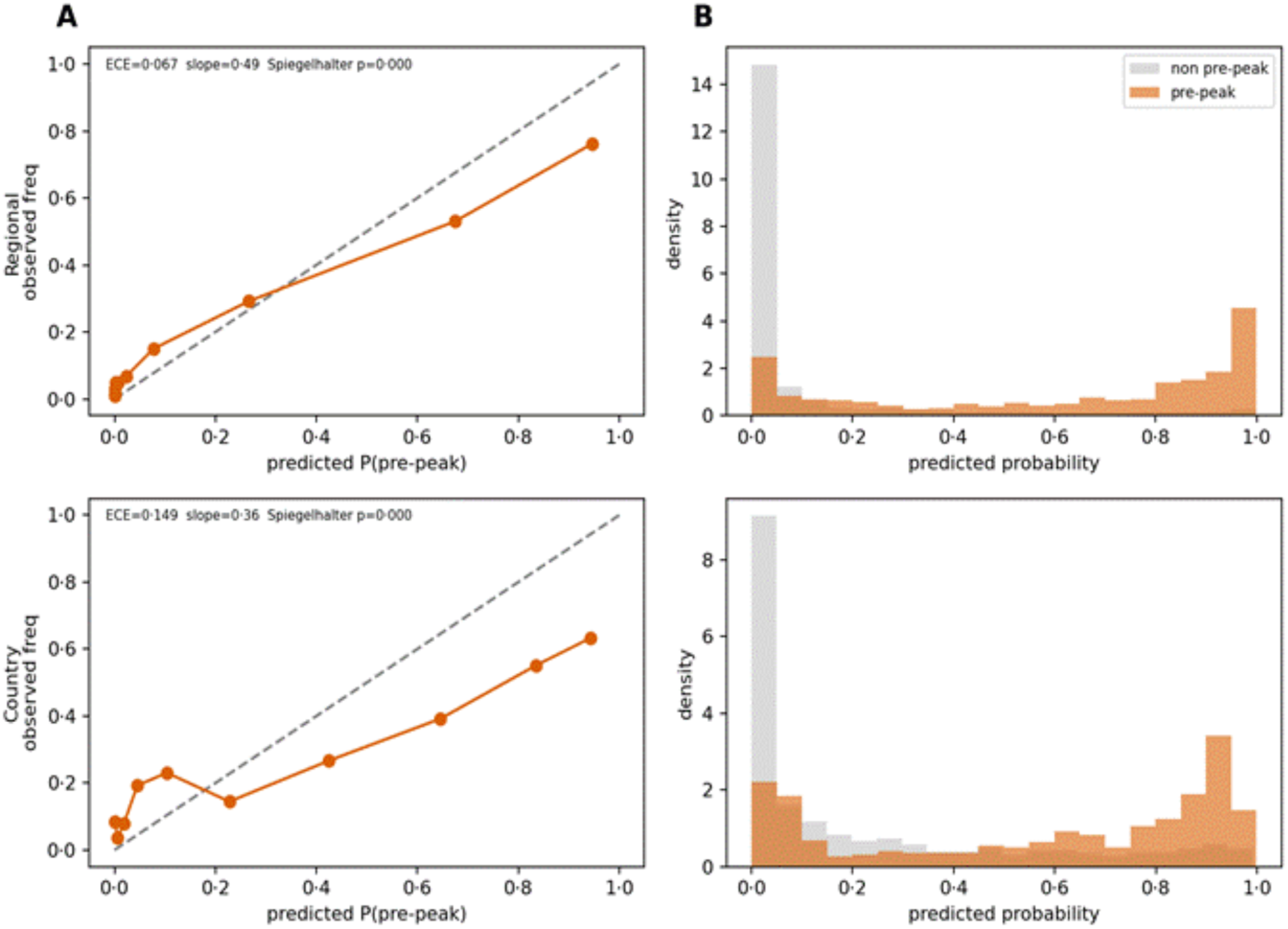
Risk-score separation and calibration by scale. Reliability curves show observed epidemic onset-phase frequency against the predicted onset-risk score, and histograms show score distributions for epidemic onset-phase and non-onset weeks. The regional panel represents the deployment scale for dengue anticipatory action and shows clear separation between low-risk and high-risk weeks. The country panel tests transportability across heterogeneous national surveillance systems, where a single pooled probability scale is not expected to hold. Calibration slopes below one indicate that raw scores should be locally recalibrated before use as absolute probabilities. Full statistics are in table S9.

### A rainfall rise, not a rainfall level, precedes acceleration

The attribution analysis showed that rainfall features carry seasonal timing, so we tested whether the rainfall signal precedes the onset of transmission acceleration. For each unit and within fold, we compared an interpretable accumulation level in millimetres against rainfall-dynamics representations, the rate of rise, and the rainfall short-term to long-term ratio, selecting the representation, lead, and threshold that maximised the Youden J statistic on training years and scoring the held-out year (table S8). The accumulation level was a weak detector of the pre-onset weeks at every scale, with leave-one-year-out AUROC of 0·55 across regions and 0·60 across countries. High accumulation persists throughout the wet season and does not single out the transition. The rainfall-dynamics representation was consistently stronger, reaching 0·73 across regions and 0·64 across countries (an eight-week rate of rise, with a median lead of about ten weeks). The paired dynamics-minus-accumulation difference was +0·180 across regions (0·136–0·223; p=0·002) but only +0·032 across countries (−0·086 to 0·168; p=0·622), where the difference was not significant (table S8). A sharp rainfall rise, not a high rainfall level, is therefore what anticipates acceleration.

## Discussion

Generative embedding provides the representational strategy. In its original form, generative embedding uses quantities from a generative model as features for discriminative classification.^12^ For sparse weekly dengue series, the generative step is implemented as deterministic transforms of the observed data. Each transform uses only observations available at the decision week, preserving the causal construction described in the Methods. This implementation links generative embedding with knowledge-guided machine learning and state reconstruction, where process knowledge helps represent a partially observed epidemic system.^11–13,23^ In the dengue embedding, the main features represent transmission acceleration and rainfall phase, with seasonal and susceptibility terms adding epidemic-history information.

Because the paper tests representation, catch22 serves as the generic control. It summarises temporal shape in a domain-agnostic way and tests whether transformation alone exposes predictive signal.^25^ The dengue embedding then assigns the transformed signal to epidemiological features, separating rainfall rise from accumulation and acceleration from generic temporal shape. These features map to established components of dengue emergence, including hydrometeorological forcing, temporal dependence near transitions, and susceptible turnover across epidemic cycles.^14–16,22,23^ The evaluated epidemic onset is defined from the case series by the transmission-acceleration framework, consistent with surveillance-based outbreak detection.^17–19^ The analysis therefore tests causal reconstruction of that onset using information available at the decision week.

Spatial variation tests where that reconstruction remains coherent. A rainfall-paced embedding requires alignment between rainfall phase and epidemic timing within the analytical unit. Regional surveillance units preserve this alignment more directly than national series, which can pool distinct climate zones and asynchronous epidemic phases.^14,28^ National aggregation can attenuate the climate-paced signal represented by the embedding. For this reason, regional risk scoring is the relevant calibration scale.^26^

Because the target is acceleration-defined onset, acceleration-derived features should be read as features tied to that definition. The retrospective validation further restricts interpretation to seasons with a qualifying onset; national estimates are less stable when aggregation leaves few coherent seasons. Prospective deployment requires reporting-delay correction, local recalibration, and threshold evaluation before operational use. For sparse epidemiological time series, from neglected infectious diseases to other climate-sensitive infections, useful foundation-model early warning is likely to depend on generative embeddings of the variables already collected in surveillance.

## Use of artificial intelligence

Generative artificial-intelligence tools were used during manuscript preparation for copy-editing, structural review, and critique of author-drafted text. The tools were Claude Opus 4.6 (Anthropic) and GPT-4 (OpenAI). No artificial-intelligence tool ran the analytical pipeline, chose features or thresholds, produced empirical results, selected citations, or determined the final interpretation. All tool-assisted revisions were reviewed, edited, and approved by the authors, who retained responsibility for the final wording and all scientific claims. No artificial-intelligence tool is listed or cited as an author.

## Data Availability

All data produced are available online at the Humanitarian Data Exchange.

https://github.com/pelitro-rcw/dengue-tabular-models

## Contributors

DB conceptualised the study and designed the reconstruction and evaluation. AML secured funding. TOM, GMG, JFM, KS, KB, and AJDA conducted data collection and curation. DB and KJP implemented the analytical pipeline and performed the analysis. DB and AML supervised the study. All authors interpreted the results and contributed to the manuscript. DB and KJP accessed and verified the underlying data. All authors had access to the data, approved the final version, and accepted responsibility for the decision to submit.

## Data sharing

All data needed to reproduce the analysis are available. The data analysed are the deidentified weekly dengue case counts for the regional and national scales, together with the area-weighted weekly rainfall series; these come from the same public repositories.^19^ The regional Philippine panel is available from the Humanitarian Data Exchange and the associated deposited dataset, and the eight-country panel is available from the OpenDengue repository.^20^ Additional materials accompany the deposit: a data dictionary defining each field, the enriched feature matrix, the feature dictionary, and a reproducibility manifest documenting execution order and expected outputs. The full analytical code for the reconstruction, the evaluation, and the figures is deposited at GitHub under an MIT licence (https://github.com/pelitro-rcw/dengue-tabular-models). The data and code are available on publication and carry no access restriction or embargo; no proposal, data-access agreement, or investigator support is required for re-use. Persistent identifiers and the public code-repository link will be provided on acceptance.

## Acknowledgements

We thank the Philippine Department of Health Epidemiology Bureau and the Humanitarian Data Exchange for the regional Philippine dataset. This work was funded by the National Institute of Environmental Health Sciences of the National Institutes of Health (Award Number P20ES036118). The content is solely the responsibility of the authors and does not necessarily represent the official views of the National Institutes of Health.

## Appendix

**Table S1:**
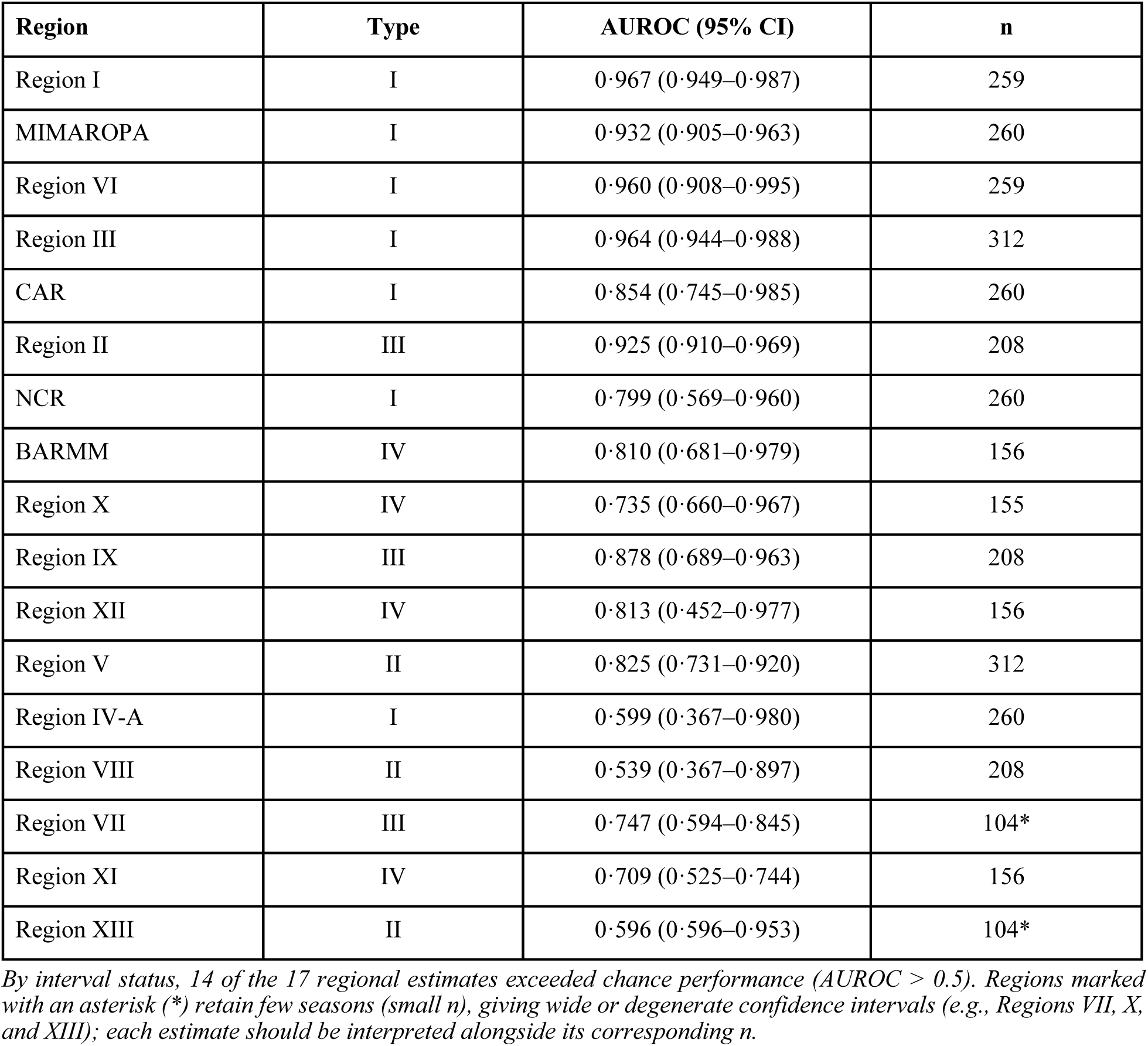
Per-region detection at the regional scale (Generative Embedding + TabPFN, leave-one-year-out), by PAGASA climate type.

| Region | Type | AUROC (95% CI) | n |
| --- | --- | --- | --- |
| Region I | I | 0.967 (0.949–0.987) | 259 |
| MIMAROPA | I | 0.932 (0.905–0.963) | 260 |
| Region VI | I | 0.960 (0.908–0.995) | 259 |
| Region III | I | 0.964 (0.944–0.988) | 312 |
| CAR | I | 0.854 (0.745–0.985) | 260 |
| Region II | III | 0.925 (0.910–0.969) | 208 |
| NCR | I | 0.799 (0.569–0.960) | 260 |
| BARMM | IV | 0.810 (0.681–0.979) | 156 |
| Region X | IV | 0.735 (0.660–0.967) | 155 |
| Region IX | III | 0.878 (0.689–0.963) | 208 |
| Region XII | IV | 0.813 (0.452–0.977) | 156 |
| Region V | II | 0.825 (0.731–0.920) | 312 |
| Region IV-A | I | 0.599 (0.367–0.980) | 260 |
| Region VIII | II | 0.539 (0.367–0.897) | 208 |
| Region VII | III | 0.747 (0.594–0.845) | 104* |
| Region XI | IV | 0.709 (0.525–0.744) | 156 |
| Region XIII | II | 0.596 (0.596–0.953) | 104* |

*By interval status, 14 of the 17 regional estimates exceeded chance performance (AUROC > 0.5). Regions marked with an asterisk (\*) retain few seasons (small n), giving wide or degenerate confidence intervals (e.g., Regions VII, X, and XIII); each estimate should be interpreted alongside its corresponding n.*

**Table S2:** Generic-feature control: domain-only versus domain-plus-canonical (catch22) (leave-one-year-out). The AUROC gain (enriched minus domain-only) carries a 95% cluster-bootstrap interval and a paired p value.

| Scale | Domain-only AUROC | Enriched AUROC | Enriched – domain ΔAUROC (95% CI; p) |
| --- | --- | --- | --- |
| Regional | 0.881 | 0.887 | +0.006 (–0.005 to 0.017; p = 0.304) |
| Country | 0.757 | 0.767 | +0.010 (–0.034 to 0.056; p = 0.676) |

*The generic catch22 features add little additional discrimination beyond the domain reconstruction at either scale, with small non-significant positives regionally and nationally. The domain embedding is retained because it supplies epidemiological interpretability, while catch22 remains the generic-shape control.*

**Table S3:** Per-unit rainfall trigger preceding transmission-acceleration onset (leave-one-year-out). Accumulation level (mm) versus the best rainfall-dynamics representation; † marks units whose accumulation interval excludes 0·5.

| Unit | Accum. threshold | Accum. AUROC (95% CI) | Best dynamics representation | Dyn. AUROC | Lead (wk) |
| --- | --- | --- | --- | --- | --- |
| Regional (median) | 211.5 mm | 0.514 (0.375–0.691) | Rainfall STA/LTA (3/12 wk) | 0.750 | 10.0 |
| NCR | 155.5 mm | 0.714 (0.618–0.798) † | Rainfall STA/LTA (4/26 wk) | 0.922 | 6.0 |
| Region I | 60.9 mm | 0.650 (0.561–0.731) † | Rainfall STA/LTA (4/26 wk) | 0.909 | 7.0 |
| Region III | 133.4 mm | 0.618 (0.558–0.683) † | 8-week rate of rise | 0.806 | 7.0 |
| Region X | 264.1 mm | 0.711 (0.583–0.846) † | Rainfall STA/LTA (4/26 wk) | 0.694 | 11.0 |
| Region XIII | 792.3 mm | 0.940 (0.917–0.972) † | 8-week rate of rise | 0.989 | 6.5 |
| Country (median) | 194.9 mm | 0.615 (0.415–0.765) | 8-week rate of rise | 0.672 | 10.5 |
| Brazil | 193.6 mm | 0.901 (0.770–0.946) † | 8-week rate of rise | 0.917 | 2.0 |
| Mexico | 18.5 mm | 0.615 (0.497–0.765) | Rainfall STA/LTA (3/12 wk) | 0.863 | 13.0 |
| Taiwan | 196.2 mm | 0.707 (0.540–0.783) † | 8-week rate of rise | 0.878 | 7.0 |

*Selected units shown (representative of 17 regions + 8 countries). Dynamics out-discriminate accumulation in most units. Peru is not estimable at the country scale. Detection is strongest in seasonal systems and weakest where rainfall is year-round or coastally opposing.*

**Table S4:** Feature families that reconstruct the latent state from the two raw columns.

| Class group | n | Representative features | Domain mechanism |
| --- | --- | --- | --- |
| Lags | 18 | case_lag1–16; rain_lag1–16 | recent transmission; delayed rainfall effect |
| Rolling mean, sum, SD | 21 | case_mean2–26; rain_sum4–26 | smoothed level and volatility |
| Rainfall accumulation | 6 | rain_accum4–16 | habitat persistence and breeding threshold |
| Transmission acceleration | 3 | case_stalta; stalta_delta | onset of epidemic growth (STA/LTA ratio) |
| Momentum | 7 | case_delta; case_accel; case_pct | rate and acceleration of change |
| Week-of-year climatology | 10 | case_woy_anom; weeks_into_year | anomaly versus prior-year season |
| Fourier seasonality | 6 | sin1–3; cos1–3 | explicit annual periodicity |
| Phase-transition views | 5 | rain_logistic_Rc250–550; rain_hill_R8 | threshold and saturation of rainfall |
| Low-frequency climate | 8 | rain_long52_mean; rain_cumyear; ix_enso_proxy | ENSO-like interannual forcing |
| Critical slowing down | 4 | case_var12; case_ac1_12 | rising variance and autocorrelation |
| Susceptibility reconstruction/immunity | 3 | inter_epidemic_interval; case_sum52; susceptibility_proxy | susceptibility clock and immunity depletion |
| Interactions | 4 | rain8_x_caseStalta; rain8_x_cos1 | climate by season and by state |
| Transforms and raw | 13 | log_case; case_max12; case_min12 | compression, extrema, and the originals |
| Generic canonical (catch22) | 24 | c22fb_recent_over_old; c22fb_slope | off-the-shelf time-series shape (control) |

**Table S5:** Enrichment ladder: AUROC as feature families are added cumulatively (leave-one-year-out).

| Rung | Regional | Country |
| --- | --- | --- |
| Raw two columns | 0.704 | 0.562 |
| + multi-scale case views | 0.840 | 0.732 |
| + climate and rainfall accumulation | 0.867 | 0.796 |
| + transmission biology (domain-only) | 0.881 | 0.757 |
| + generic canonical (catch22) | 0.887 | 0.767 |

*At the regional scale the final rung adds only slightly beyond the domain-only reconstruction (0.881 to 0.887). At the country scale, the climate-rainfall rung is highest (0.796), while the prespecified full enriched representation remains above raw inputs and adds little beyond the domain-only reconstruction (0.757 to 0.767). Formal tests are in table S2.*

**Table S6:**
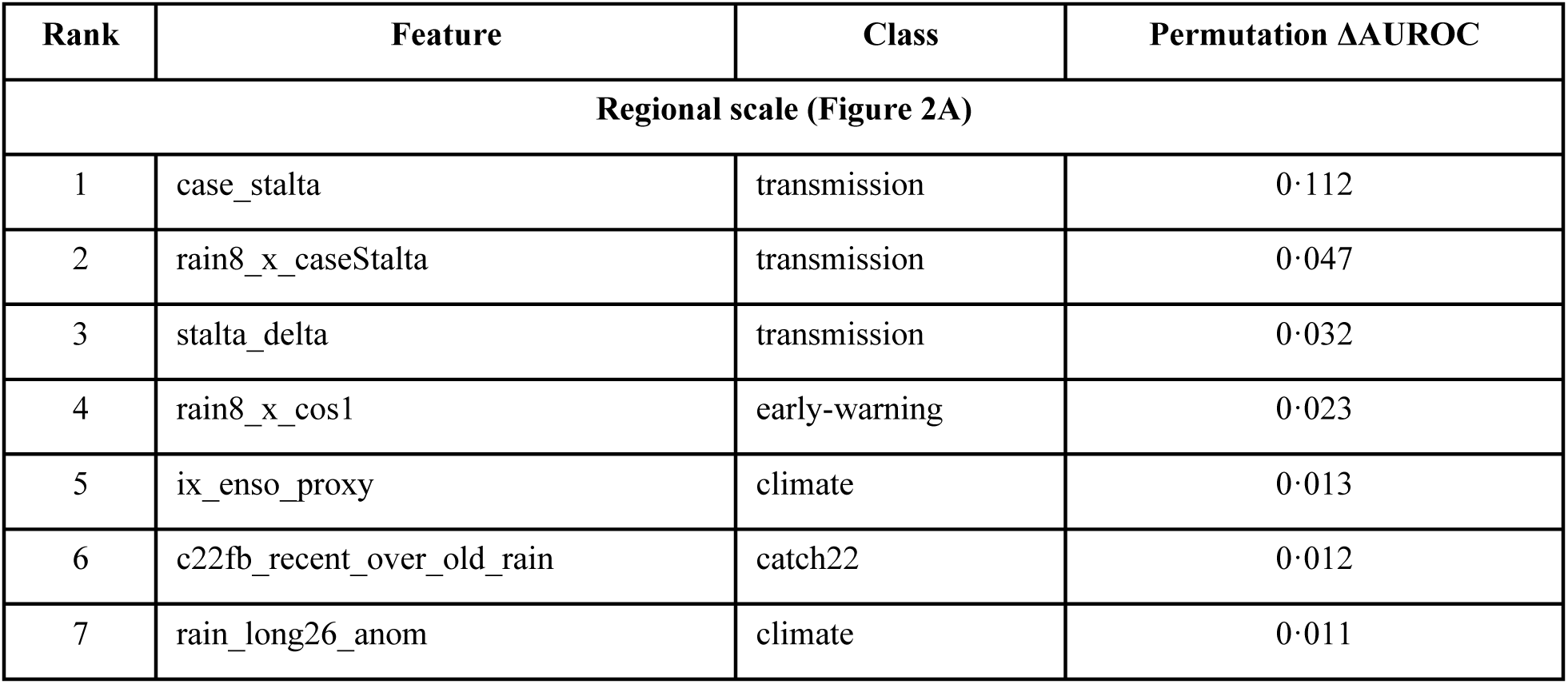
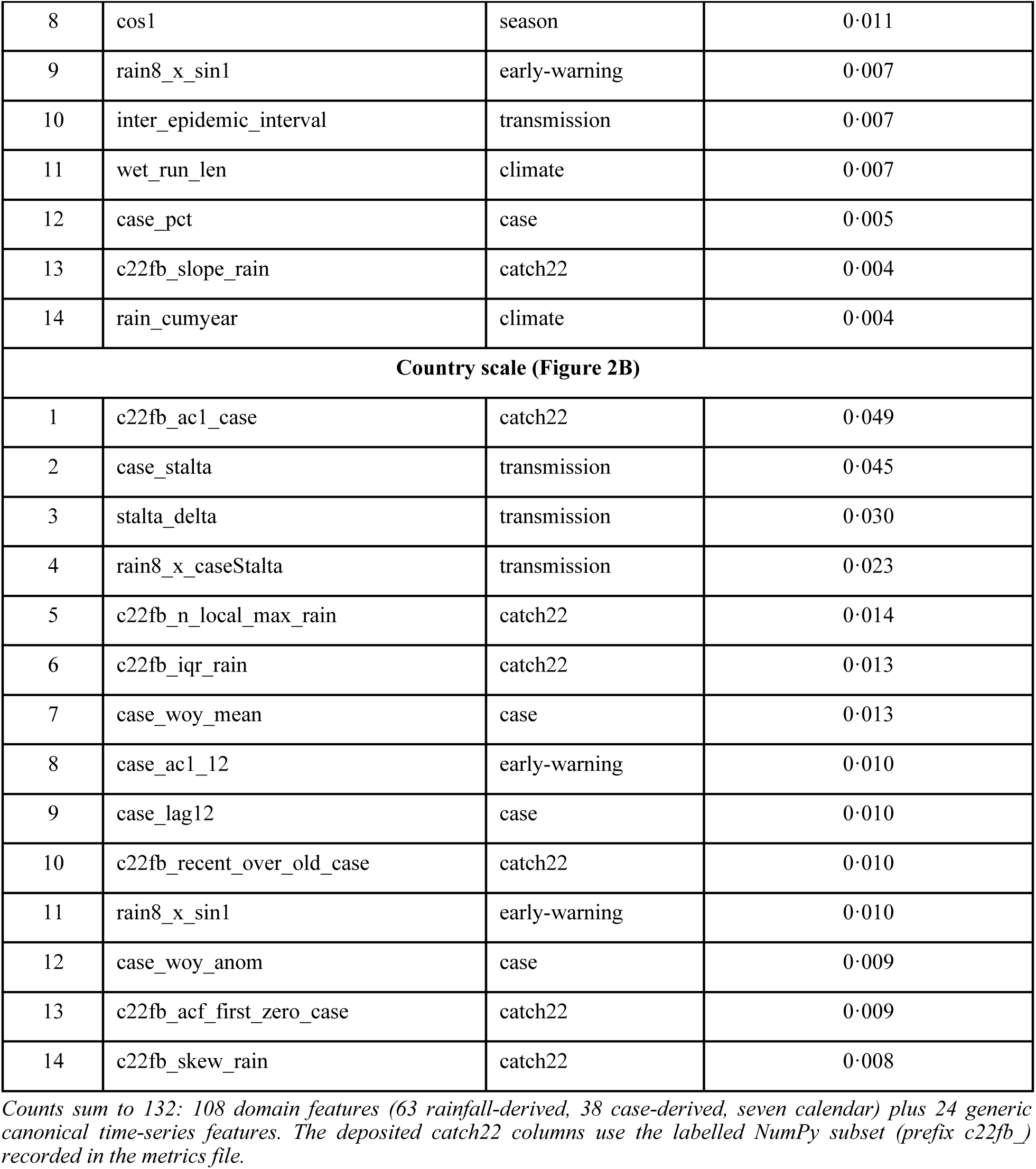
Feature attribution by scale. Leave-one-year-out permutation importance (the loss in AUROC when a feature is randomly permuted) for the 14 most predictive features at each scale, ranked; these are the values shown in Figure 2 (panel A, regional; panel B, country).

**Table S7:** Regional predictability by climate type (mean AUROC across regions in each type).

| Climate type | Rainfall regime | Regions (n) | Mean AUROC |
| --- | --- | --- | --- |
| Type I | Two pronounced seasons; dry November to April | 7 | 0·868 |
| Type II | No dry season; pronounced rainfall maximum | 3 | 0·654 |
| Type III | Short or not very pronounced dry season | 3 | 0·850 |
| Type IV | Rainfall more or less evenly distributed | 4 | 0·767 |

*Climate types follow the published Philippine climate zonation. Detection is highest in the strongly seasonal Type I regions. The across-type difference is not significant at $\alpha = 0\cdot05$ (Kruskal-Wallis $H = 5\cdot99$ , $p = 0\cdot112$ ; $n = 17$ regions). Per-region values are in table S1.*

**Table S8:** Rainfall trigger preceding the onset of transmission acceleration, by scale (leave-one-year-out). An interpretable rainfall-accumulation level is compared against the best rainfall-dynamics representation; the difference is paired and cluster-bootstrapped.

| Scale | Accumulation AUROC | Dynamics AUROC | Dynamics – accumulation (95% CI), p |
| --- | --- | --- | --- |
| Regional | 0·547 | 0·727 | +0·180 (0·136 to 0·223), $p = 0\cdot002$ |
| Country | 0·604 | 0·636 | +0·032 (–0·086 to 0·168), $p = 0\cdot622$ |

**Table S9:**
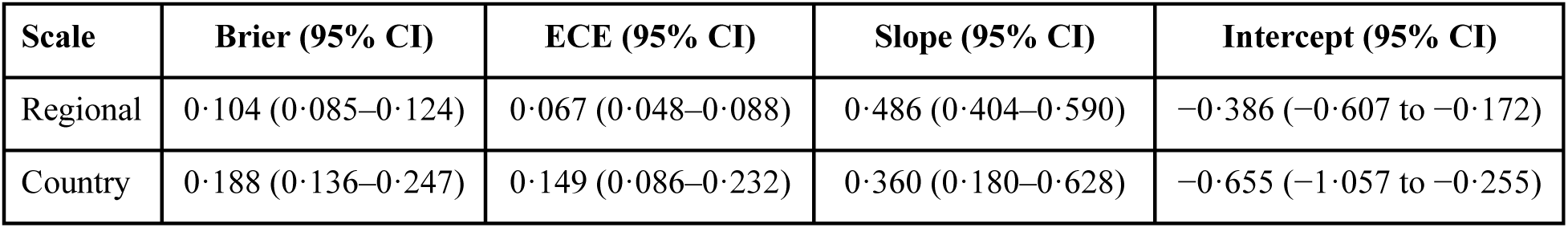
Calibration of the Generative Embedding model, by scale (leave-one-year-out). Brier, expected calibration error (ECE), and the Cox calibration slope and intercept, each with 95% cluster-bootstrap intervals.

| Scale | Brier (95% CI) | ECE (95% CI) | Slope (95% CI) | Intercept (95% CI) |
| --- | --- | --- | --- | --- |
| Regional | 0·104 (0·085–0·124) | 0·067 (0·048–0·088) | 0·486 (0·404–0·590) | −0·386 (−0·607 to −0·172) |
| Country | 0·188 (0·136–0·247) | 0·149 (0·086–0·232) | 0·360 (0·180–0·628) | −0·655 (−1·057 to −0·255) |

